# Disability evaluation in patients with Guillain-Barre syndrome and SARS-CoV-2 infection from a neurological reference center in Peru

**DOI:** 10.1101/2023.02.22.23286287

**Authors:** Sofia Stefanie Sanchez Boluarte, Wilfor Aguirre Quispe, Arantxa Noelia Sanchez Boluarte, Jhon Tacunan Cuellar, Darwin Alberto Segura Chávez

## Abstract

**Introduction:** several cases of Guillain-Barre Syndrome (GBS) associated with SARS-CoV-2 infection have been described. This study illustrated the demographic, clinical, and neurophysiological characteristics of patients with GBS and COVID-19, as well as associated factors with disability at discharge.

**Methods:** A retrospective analytical observational study was conducted. It included patients diagnosed with GBS admitted in a national reference center in Peru between 2019 and 2021. Epidemiological, clinical, neurophysiological and cerebrospinal fluid data were analyzed. A multivariate analysis, using the generalized linear model, was performed, considering the presence of disability at discharge as the dependent variable.

**Results:** 81 subjects diagnosed with GBS were included. The mean age was 46.8 years (SD: 15.2), with a predominance of males (61.73%). The most frequent clinical presentation was the classic sensory-motor form in 74 cases (91.36%) with AIDP (82.35%) as the most frequent neurophysiological pattern in the group with COVID-19, while AMAN pattern predominated (59.26%) in those without COVID-19 (p=<0.000). The disability prevalence ratio at discharge between subjects with COVID-19 and those without COVID-19 was 1.89 (CI 1.06–3.34), p=0.030, adjusted for age, sex, and neurophysiological subtype.

**Conclusions:** The neurophysiologic subtype AIDP, and a higher disability were associated with the presence of COVID-19.

## Introduction

Coronavirus disease that first appeared in China in December 2019 is currently a pandemic with over two hundred million cases and over four million deaths (1). SARS-CoV-2 infection is a multi-systemic disorder that manifests primarily at the respiratory level causing respiratory failure in severe cases and triggering aa multisystem response(2–4). Currently, many neurological manifestations associated with SARS-CoV-2 infection have been reported, including Guillain-Barre syndrome (GBS) (4–6).

GBS is an immune mediated acute inflammatory polyneuropathy, preceded by a respiratory or gastrointestinal infection in 70% of cases which typically manifests as a progressive, areflexic, ascending quadriparesis. However, other variants have also been described. Based on electrophysiological and pathological characteristics, GBS has been classified as acute inflammatory demyelinating polyradiculoneuropathy (AIDP), acute motor axonal neuropathy (AMAN) and acute sensory motor axonal neuropathy (AMSAN) (7–10).

Previous epidemics have been associated with outbreaks of GBS, describing its association with several agents such as *Campylobacter jejun*i, *Haemophilus influenzae, Epstein-barr, Cytomegalovirus, Zika*, among others (11,12). GBS is one of the most frequently associated diseases of the peripheral nervous system with SARS-CoV-2 infection (13). This study described demographic characteristics, clinical and neurophysiological manifestations of patients with GBS and SARS-CoV-2, and factors associated with disability at hospital discharge.

## METHODS

We performed a retrospective analytical observational study approved by the Institutional Review Board of the National Institute of Neurological Sciences (INCN), Lima, Peru. We included patients with the diagnosis of GBS, admitted at the INCN, between 2018 and 2021. The INCN is the national reference center for neurological diseases in Peru where only moderate to severe cases of GBS are hospitalized.

During the COVID-19 pandemic, patients with GBS continued to be admitted to our center. However, those with severe respiratory symptoms were referred to general hospitals where COVID-19 cases were treated. The study included patients above 18 years of age, diagnosed with GBS according to the Brighton criteria. COVID-19 was diagnosed using the polymerase chain reaction (PCR) test for SARS-CoV-2 based on nasopharyngeal swab detection or by detecting the anti-SARS-CoV-2 antibodies.

Conditions that mimic GBS such as acute muscle or peripheral nerve diseases were excluded from the study. Medical records were used to collect epidemiological, clinical, and laboratory data including age, sex, duration of the disease, cerebrospinal fluid (CSF) characteristics, electromyography, medical history and management.

The patients underwent one electromyography after the first week of symptoms, and the neurophysiological diagnosis was obtained according to the Uncini’s criteria (Uncini 2018). Disability was measured using the Hughes functional graduation scale as follows: 0, healthy; 1, minor symptoms, able to run; 2, able to walk 10 meters to more without support, unable to run; 3, able to walk 10 meters with help; 4, prostrate or wheelchair; 5, requires assisted ventilation; 6: dead(14). Disability was measured at admission, nadir (the day the highest score was reached) and at discharge, defined as a score ≥3 on the Hughes Scale. Also, the treatment response was defined as the decrease by at least one point on the Hughes Discharge Scale. Sample size was not considered in the study since all patients diagnosed with GBS during the reported period were included.

The mean or median, standard deviation, interquartile range (IQR) were used for continuous variables, and the categorical variables were expressed with frequencies and percentages. Normal assessment was performed using Shapiro Wilk test and the analysis between the groups was performed using a t-test in cases of quantitative variables with normal distribution or using Mann–Whitney U test. Additionally, chi-2 test and Fisher’s exact test were used for categorical variables. The statistical significance was set at 0.05. The groups were compared based on COVID-19 diagnosis, pre- and post-pandemic periods, and disability at discharge. Likewise, a generalized linear regression model was constructed, with the presence of disability at discharge as a dependent variable. The analysis was performed using STATA (v.17.0, Texas, USA).

## RESULTS

We included 81 patients diagnosed with GBS between January 2018 and April 2021. The mean age of the patients was 46.8 years (SD: 15.2), with a predominance of the male sex (61.73%). Forty-four (54.32%) cases met level 1 of diagnostic certainty, 26 (32.10%) cases met level 2 and 11 (13.58%) met level 3. The median time of disease at admission was 5 days (IQR: 3.0 -8.0). The mean of the Hughes scale at both admission and nadir was 3.5(SD:0.9), with the time at nadir of 7 days (IQR: 4.0-10.0). The most frequent clinical presentation was the classic motor sensory form in 74 cases (91.36%), 4 (4.94%) presented with Miller-Fisher syndrome, 2 (2.47%) with Bickerstaff encephalitis, and 1 (1.23%) patient with facial diparesis.

Thirty-two (50.79%) subjects had a history of gastrointestinal infection and 17 (26.98%) with a history of respiratory infection. The CSF analysis showed albumin cytological dissociation in 49 (63.64%) patients. Electromyography was performed on 71 participants, of whom 39 (48.15%) underwent EMG after the 4^th^week of symptoms. Thirty-three (46.48%) patients had AMAN, 29 (40.85%) AIDP, 4 (5.63%) AMSAN and 5 (7.05%) presented with no alterations in the study. The median hospitalization time was 14 days (IQR:9.0-21.0). The median for Hughes at discharge was 3 days (IQR: 2.0 - 3.0). Nine (14.06%) required mechanical ventilation. The results of treatment are as follows, 67 (83.75%) received intravenous immunoglobulin, 6 (7.50%) received plasma replacement, 41 (50.62%) had response to treatment (Table 1). From the period of March 2020 to April 2021, 31 subjects (38.3%) were diagnosed as GBS, of which 18 (58.0%) had a diagnosis of COVID-19, while in the period before the pandemic, from January 2019 to February 2020, 50 subjects (61.7%) received the diagnosis of GBS.

**Table 1.**
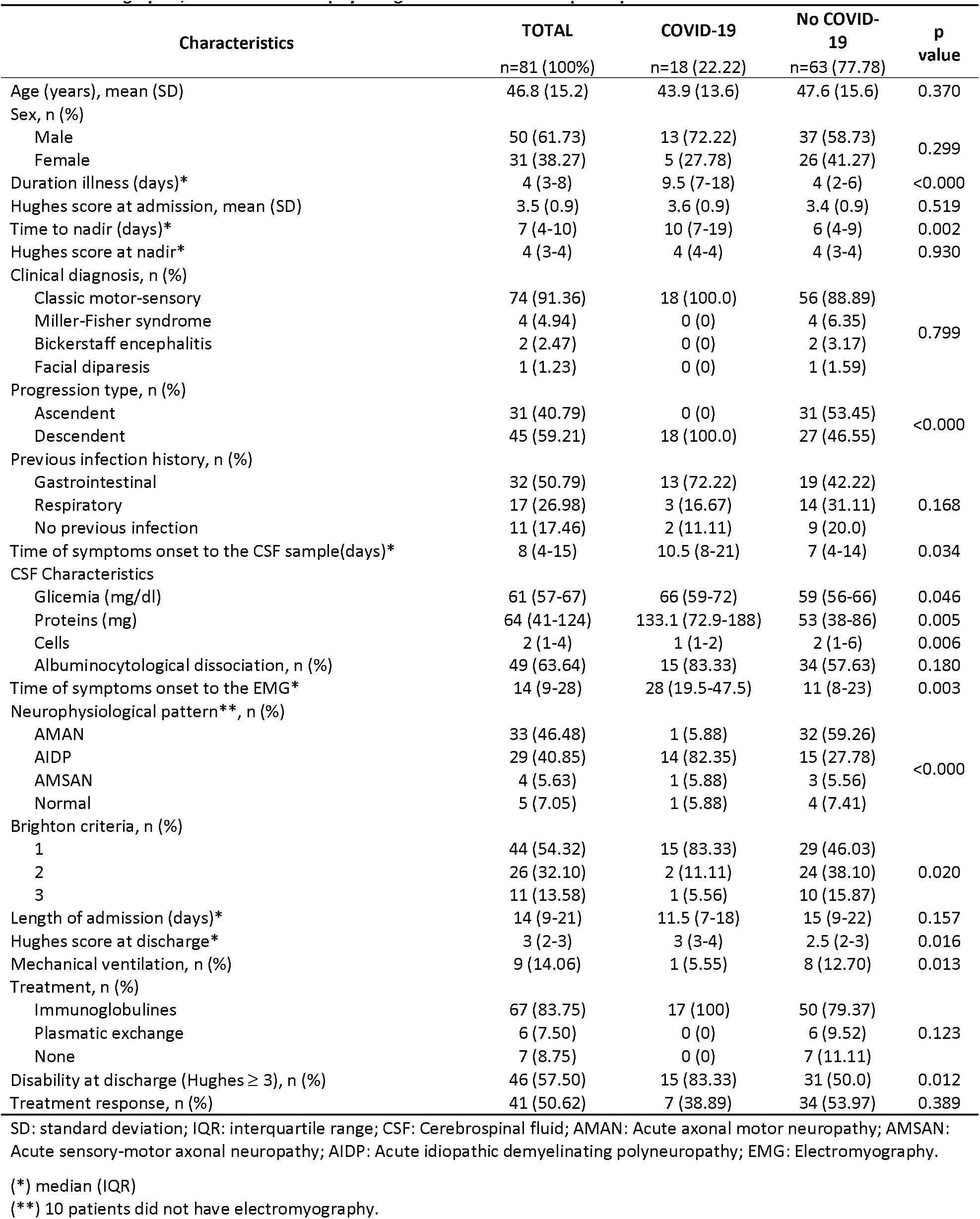
Demographic, clinical and neurophysiological characteristics of participants in relation to SARS-CoV-2 infection.

**Table 2.** Characteristics of patients according to their state of disability at hospital discharge.

| Characteristics | Disability<br>n=46 (57.50%) | No disability<br>n=34 (42.50%) | P value |
| --- | --- | --- | --- |
| Age (years), mean (SD) | 48.4 (15.4) | 44.2 (14.7) | 0.225 |
| Sex, n (%) |  |  |  |
| Male | 28 (60.87) | 22 (64.71) | 0.726 |
| Female | 18 (39.13) | 12 (35.29) |  |
| Duration illness (days)* | 5 (2-9) | 4 (3-7) | 0.477 |
| Hughes score at admission, mean (SD) | 4 (3-4) | 3.1 (0.9) | 0.002 |
| Time to nadir (days)* | 7 (4-11) | 5 (4-9) | 0.131 |
| Hughes score at nadir* | 4 (4-4) | 3.5 (2-4) | <0.000 |
| Clinical diagnosis, n (%) |  |  |  |
| Classic motor-sensory | 44 (95.65) | 29 (85.29) |  |
| Miller-Fisher syndrome | 0 (0) | 4 (11.76) | 0.009 |
| Bickerstaff encephalitis | 2 (4.35) | 0 (0) |  |
| Facial diparesis | 0 (0) | 1 (2.94) |  |
| Progression type, n (%) |  |  |  |
| Ascendent | 16 (34.78) | 14 (48.28) | 0.245 |
| Descendent | 30 (65.22) | 15 (51.72) |  |
| Previous infection history, n (%) |  |  |  |
| Gastrointestinal | 19 (50.0) | 13 (54.17) |  |
| Respiratory | 9 (23.68) | 7 (29.17) | 0.663 |
| No previous infection | 7 (18.42) | 4 (16.67) |  |
| CSF Characteristics* |  |  |  |
| Glicemia (mg/dl) | 61 (58-67) | 59.5 (56-70) | 0.464 |
| Proteins (mg) | 64 (39-112) | 67 (41-133) | 0.627 |
| Cells | 2 (1-3) | 2 (1-8) | 0.122 |
| Albuminocytological dissociation, n (%) | 27 (67.50) | 22 (78.57) | 0.317 |
| Neurophysiological pattern *, n (%) |  |  |  |
| AMAN | 17 (41.46) | 16 (53.33) |  |
| AIDP | 20 (48.78) | 9 (30.0) | 0.429 |
| AMSAN | 2 (4.88) | 2 (6.67) |  |
| Normal | 2 (4.88) | 3 (10.0) |  |
| Brighton criteria, n (%) |  |  |  |
| 1 | 27 (58.70) | 17 (50.0) |  |
| 2 | 13 (28.26) | 13 (38.24) | 0.640 |
| 3 | 6 (13.04) | 4 (11.76) |  |
| Length of admission (days)* | 15.5 (10-23) | 13 (8-17) |  |
| Mechanical ventilation, n (%) | 7 (15.22) | 1 (2.94) | 0.070 |
| Time to the treatment onset (days)* | 6 (3-9) | 6 (5-11.5) | 0.289 |
| Treatment, n (%) |  |  |  |
| Immunoglobulines | 40 (88.89) | 26 (76.47) |  |
| Plasmatic exchange | 2 (4.44) | 3 (8.82) | 0.249 |
| None | 3 (6.67) | 5 (14.71) |  |
| Treatment response, n (%) | 16 (34.78) | 25 (73.53) | 0.003 |

SD: standard deviation; IQR: interquartile range; CSF: Cerebrospinal fluid; AMAN: Acute axonal motor neuropathy; AMSAN: Acute sensory-motor axonal neuropathy; AIDP: Acute idiopathic demyelinating polyneuropathy; EMG: Electromyography.
(\*) 10 patients did not have electromyography.

Patients during the pandemic period had longer duration of illness when compared to the groups treated before the pandemic (2019-february 2020), 7 (IQR 4.0–10.0) vs 4 (IQR 2.0–6.0), p=0.006; and a longer time at nadir, 9.0 (IQR 5.0–15.0) vs 5.5 (IQR 3.0–8.0), p=0.002; and a shorter hospitalization time, 12 days (IQR 7.0–18.0) vs 16 days (IQR 9.0–26.0), p=0.033. Twenty-nine (83.8%) received intravenous immunoglobulin during the pandemic, whereas only 38 (76.0%), p=0.046 before the pandemic. There was neither a difference in Hughes’ score at admission, nor in the response rate to the treatment comparing both periods. (Table 3)

**Table 3.** Demographic, clinical, and neurophysiological characteristics of GBS cases according to pre- and post-pandemic period.

| Characteristics | January 2019 - February 2020<br>n=50 (61.73%) | March 2020 - April 2021<br>n=31 (38.27%) | P<br>value |
| --- | --- | --- | --- |
| Age (years), mean (SD) | 48 (16.3) | 44.8 (13.1) | 0.365 |
| Sex, n (%) |  |  |  |
| Male | 19 (38.0) | 12 (38.7) | 0.949 |
| Female | 31 (62.0) | 19 (61.3) |  |
| Duration illness (days)* | 4 (2-6) | 7 (4-10) | 0.006 |
| Hughes score at admission, mean (SD) | 4 (3-4) | 4 (3-4) | 0.4257 |
| Time to nadir (days)* | 5.5 (3-8) | 9 (5-15) | 0.002 |
| Hughes score at nadir* | 4 (3-4) | 4 (3-4) | 0.474 |
| Clinical diagnosis, n (%) |  |  |  |
| Classic motor-sensory | 45 (90.0) | 29 (93.6) | 0.79 |
| Miller-Fisher syndrome | 2 (4.0) | 2 (6.5) |  |
| Bickerstaff encephalitis | 2 (4.0) | 0 |  |
| Pharyngocervicobrachial | 0 | 0 |  |
| Facial diparesis | 1 (2.0) | 0 |  |
| Progression type, n (%) |  |  |  |
| Ascending | 23 (48.9) | 8 (27.6) | 0.066 |
| Descending | 24 (51.1) | 21 (72.4) |  |
| Previous infection history, n (%) |  |  |  |
| Gastrointestinal | 16 (43.2) | 16 (61.5) | 0.243 |
| Respiratory | 12 (32.4) | 5 (19.2) |  |
| No previous infection | 6 (16.22) | 5 (19.2) |  |
| Time of symptoms onset to the CSF sample* | 9 (4-15) | 8 (5-13) | 0.497 |
| CSF Characteristics* |  |  |  |
| Glicemia (mg/dl) | 59 (55-64) | 64.5 (59-72) | 0.006 |
| Proteins (mg) | 54 (39-86) | 83 (42-153) | 0.152 |
| Cells | 2.5 (1-8) | 1 (1-2) | 0.001 |
| Albuminocytological dissociation, n (%) | 26 (56.5) | 23 (74.2) | 0.036 |
| Time of symptoms onset to the EMG* | 11 (8-17) | 22 (14-33) | 0.009 |
| Neurophysiological pattern *, n (%) |  |  |  |
| AMAN | 27 (61.4) | 6 (22.2) | 0.007 |
| AIDP | 11 (25.0) | 18 (66.7) |  |
| AMSAN | 2 (4.6) | 2 (7.4) |  |
| Normal | 2 (4.6) | 1 (3.7) |  |
| Brighton criteria, n (%) |  |  |  |
| 1 | 22 (44.0) | 22 (71.0) | 0.059 |
| 2 | 20 (40.0) | 6 (19.4) |  |
| 3 | 8 (16.0) | 3 (9.7) |  |
| Length of admission (days)* | 16 (9-26) | 12 (7-18) | 0.033 |
| Hughes score at discharge, n (%) | 3 (2-3) | 3 (2-4) | 0.212 |
| Mechanical ventilation, n (%) | 7 (14.0) | 2 (14.3) | 0.978 |
| Time to the treatment onset (days)* | 6 (4-9) | 5 (5-10) | 0.582 |
| Treatment, n (%) |  |  |  |
| Immunoglobulines | 38 (76.0) | 29 (83.8) | 0.046 |
| Plasmatic exchange | 6 (12.0) | 1 (3.3) |  |
| None | 6 (12.0) | 0 |  |
| Disability at discharge (Hughes ≥ 3), n (%) | 26 (53.1) | 20 (64.5) | 0.313 |
| Treatment response, n (%) | 27 (55.1) | 15 (50.0) | 0.659 |

SD: standard deviation; IQR: interquartile range; CSF: Cerebrospinal fluid; AMAN: Acute axonal motor neuropathy; AMSAN: Acute sensory-motor axonal neuropathy; AIDP: Acute idiopathic demyelinating polyneuropathy; EMG: Electromyography.
(\*) median (IQR)
(\*\*) 10 patients did not have electromyography.

**Table 4.** Associated factors for epilepsy on the multiple regression analysis.

| Variables | Bivariate analysis |  |  | Multiple regression* |  |  |
| --- | --- | --- | --- | --- | --- | --- |
|  | OR | IC 95% | p | OR | IC 95% | p |
| Sex |  |  |  |  |  |  |
| Male | 0.93 | 0.64-1.37 | 0.725 | 0.74 | 0.42-1.29 | 0.286 |
| Female | Ref. |  |  | Ref. |  |  |
| Age | 1.01 | 0.99-1.02 | 0.210 | 1.01 | 0.99-1.02 | 0.117 |
| Neurophysiological diagnosis |  |  |  |  |  |  |
| Axonal | Ref. |  |  | Ref. |  |  |
| Demyelinizing | 1.34 | 0.89-2.00 | 0.149 | 1.11 | 0.63-1.94 | 0.717 |
| COVID-9 |  |  |  |  |  |  |
| No | Ref. |  |  | Ref. |  |  |
| Yes | 1.67 | 1.20-2.31 | 0.002 | 1.89 | 1.06-3.34 | 0.030 |

\*Adjusted by age, sex, and neurophysiological diagnosis.

In the period March 2020 - April 2021, 18 cases (38.3%) were detected with both COVID-19 and GBS. COVID-19 was diagnosed using PCR in 7 cases and by rapid antibody testing in 11 participants. Of the 11 cases, 6 were positive for both IgM and IgG, 1 for IgM and 2 for IgG only. Fourteen (77.8%) had respiratory symptoms. The median time between onset of respiratory symptoms and neurological symptoms was 9 days (IQR:6.0-13.0). The COVID-19 group had thirteen (72.2%) males whereas 37 (58.73%) males were present in the group without COVID-19, p=0.299. Mean age was similar in both groups, as was the median of the Hughes scale at admission and nadir. Compared with the group without COVID-19, participants with COVID-19 had a longer time of disease at admission, 9.5 (IQR: 7.0-18.0) vs 4 (IQR: 2.0-6.0), p<0.000; and at nadir, 10 (IQR:7.0-19.0) vs 6 (IQR:4.0-9.0), p=0.002. (Table 1) The classic sensory-motor clinical presentation was seen frequently in both groups, 13 (72.22%) patients in the COVID-19 group had a history of gastrointestinal infection, and nineteen had a respiratory infection (42.2%) in the group without COVID-19. The CSF characteristics, of the patients with COVID-19 had higher protein elevation, 133.1 (IQR: 72.9 - 188.0) when compared to those without COVID-19, 53.0 (IQR 38-86), p=0.005. Albumin cytological dissociation was observed in 15 (83.3%) patients with COVID-19 vs 34 (57.63%) patients in the other group, p=0.0.180.

There were no differences in the disease duration at the time of sampling the CSF. The most frequent neurophysiological pattern in the group with COVID-19 was AIDP (82.4%), whereas AMAN predominated (59.26%), p=<0.000 in those without COVID-19. Hospitalization time was greater for those without COVID-19, 15 (IQR 9.0 - 22.0) vs 11.5 (IQR 7.0–18.0), p=0.157. Patients with COVID-19 had slightly higher score of Hughes 3 (IQR:3-4) comparing with those without COVID-19 2.5 (2-3) (p=0.016). Patients with COVID-19 had with less frequency mechanical ventilation versus those without COVID-19. (1 (5.55%) versus 8 (12.70%), p=0.013.

There was no significant difference between the Hughes at discharge, and mechanical ventilation requirement. All patients with COVID-19 received intravenous immunoglobulin at a dose of 0.4g/kg, when compared to 50 (79.37%) patients in the group without COVID-19, 6(9.52%) received plasma replacement and 7 (11.11%) did not receive treatment. Fifteen patients (83.33%) of the group with COVID-19 had disability at discharge, comparing with 31 (50.0%) on the group without COVID-19. The response rate to treatment was similar in both groups. (Table 1) Patients with disability at discharge had greater hospitalization time, greater use of mechanical ventilation and less response to treatment, when compared to the non-disabled. (Table 3) The prevalence ratio of disability at discharge in patients with and without COVID-19 was 1.89 (CI 1.06–3.34), p=0.030, adjusted for age, sex, and neurophysiological subtype.

## DISCUSSION

We found that there were no differences in the age, according to COVID-19 infection, which was unique when compared to the previously reported studies which had described a higher mean age for the group of patients with COVID-19(3). Our patients were predominantly males, with a higher proportion in the COVID-19 group, possibly due to male predominance in patients with COVID-19(15), which could predispose them to an increased risk of developing GBS.

Unlike other studies, we did not observe a significant increase in the frequency of GBS cases during the pandemic(16), due to the measures established by the Ministry of Health of Peru that led to a decrease in the flow of patients in all health facilities in the country.

Patients with GBS and COVID-19 had longer admission durations when compared to those without COVID-19, this delay in care may have been related to the longer time of isolation of patients with respiratory symptoms by COVID-19. Likewise, the time between SARS-CoV-2 infection and the onset of GBS symptoms was 9 days, which is consistent with the hypothesis of a postinfectious mechanism associated with COVID-19, rather than direct neuronal damage or a para-infectious mechanism(10). The average duration has previously been reported to be 2 weeks, like other infections associated with GBS(3,10,17).

According to the results of our study in patients with COVID-19, the clinical manifestations and distribution of clinical variants were like those of classical GBS, characterized by sensory-motor predominance and a neurophysiological AIDP pattern, similar to previous reports (17–19). Likewise, cerebrospinal fluid protein levels were higher in patients with COVID-19, as described previously (18,20) which attributes more to an immune response than to an inflammatory or infectious(18).

Independent of the concomitant infection with COVID-19, participants with GBS had a disease of similar severity both at admission and nadir; this differs from the studies reported previously, where differences have been found in the severity evaluated at admission(21), during hospitalization and at hospital discharge(17,22). However, in the multivariate analysis that resulted in disability at hospital discharge, it was determined that infection with SARS-CoV-2 is associated with greater disability, although patients admitted to our center presented with COVID-19 of mild to moderate severity. The progression of disability was found to be slower in patients with COVID-19 compared to patients without COVID-19. One reason attributed for this difference could be that most patients with COVID-19 had an electromyographic pattern corresponding to AIDP, a pattern associated with slower progression(15,23).

Although patients admitted to our center had moderate to severe GBS, none had a fatal outcome, as reported in other countries, in which no changes in mortality associated with COVID-19 were evidenced(24).

The limitations of our study included selection bias as patients with severe respiratory symptoms from COVID-19 were referred to the general hospitals for multidisciplinary management, which can influence disability-related outcomes and severity. This situation, however, was due to the restrictions on health care and hospital admission introduced during the start of the pandemic. Despite having a significant number of cases, there was no balanced ratio between the groups with and without COVID-19. The uniformity in the diagnostic criteria for SARS-CoV-2 infection was also affected because of its modification by the Peruvian Ministry of Health in the very beginning of the pandemic. Other limitation is that on our resource limited settings, we were not able to perform serial EMGs and this could introduce wrong classifications, however, we followed Uncini criteria for the diagnosis.

It was concluded that patients with COVID-19 presented more frequently with the neurophysiological subtype AIDP, an increase in the concentration of proteins in CSF. Greater disability at discharge was associated with SARS-CoV-2 infection, despite evidence of slower progression in this group. We believe that these findings can not only be useful in the diagnostic approach aimed towards patients with a recent history of COVID-19, but also as a guide to the immediate prognosis of patients in this group.

## Data Availability

All data produced in the present study are available upon reasonable request to the authors

## Acknowledgments

The authors thank Aafreen Khan for English language support.

